# Trends in medication abortion service delivery in the U.S., 2020-2025

**DOI:** 10.64898/2026.07.01.26357048

**Authors:** Shelly Kaller, Rosalyn Schroeder, Nancy F. Berglas, Clare Stewart, Ushma D. Upadhyay

## Abstract

**Objective:** Since 2020, medication abortion provision in the U.S. has been reshaped by changing abortion policies and expanded telehealth access, yet little is known about how medication abortion service delivery has evolved. We examined national trends in service delivery from 2020 to 2025, including changes in abortion facility types, telehealth provision, and gestational limits.

**Study Design:** Using ANSIRH’s Abortion Facility Database, a national census of publicly advertising abortion facilities (2020–2025), we analyzed trends in medication abortion service delivery. Systematic web searches and mystery shopper calls gathered data on facility types, telehealth provision, and gestational limits. Data analysis included frequencies and comparisons across regions and states.

**Results:** Medication abortion-only facilities increased nationally, from 35% of facilities in 2020 to 65% in 2025, with substantial growth in abortion-restrictive regions such as the Midwest and South. By 2025, 99% of facilities provided medication abortion. Telehealth provision expanded from 7 facilities in 2020 to 606 facilities by 2025, driven by growth in both brick-and-mortar facilities offering telehealth care and new virtual clinics. Overall, 46% of all facilities offered medication abortion by telehealth in 2025. Gestational limits for medication abortion increased nationally, from <1% of facilities offering medication abortion after 11 weeks in 2020 to 38% in 2025.

**Conclusions:** Medication abortion service delivery has adapted to legal and logistical challenges by increasing telehealth options and expanding gestational limits. These changes improve access for abortion seekers, especially those living in restrictive environments. Sustaining abortion access will require ongoing provider adaptation and supportive policy environments.

**Implications:** Telehealth provision, virtual clinics, and expanded gestational limits have become increasingly common features of medication abortion service delivery. These findings demonstrate how abortion facilities have adapted service delivery models during a period of substantial policy and regulatory change.

## 1. Introduction

Medication abortion is now the leading method of abortion in the U.S., growing from 53% of clinician-provided abortions, including telehealth, in 2020 to 65% in 2023 [1]. During this time, the U.S. Food and Drug Administration lifted the requirement that mifepristone be dispensed in-person, building on years of advocacy and litigation surrounding medication abortion access, allowing for an expansion of medication abortion delivery models, including through pharmacies, mobile vans, and via telehealth [2,3]. Both established brick-and-mortar abortion facilities and new online-only virtual clinics began offering medication abortion pills to patients by mail [4]. As access to in-person abortion care has become totally banned or banned at 6 weeks in 18 states since the *Dobbs v. Jackson Women’s Health Organization* decision in June 2022, including states with pre-existing bans [5], the provision of medication abortion through telehealth models has become an important new avenue for care. Telehealth provision accounted for 5% of abortions nationwide before *Dobbs* in April 2022, quickly rising to 29% three years later in December 2025 [6]. Telehealth abortion provision has continued to expand with the passing of abortion “shield laws” in several states, which provide legal protections for clinicians who provide telehealth abortions to people living in states with abortion bans or legal restrictions on telehealth abortion [6].

While prior research has documented increases in medication abortion and telehealth abortion use, less is known about how abortion service delivery infrastructure has changed during this period of rapid policy and regulatory change. Given the monumental changes to the abortion care landscape, we sought to describe how medication abortion service delivery has changed since 2020. Specifically, we examine changes in the number and types of facilities offering medication abortion and the growth of telehealth provision. We also assess how facilities’ gestational limits for providing medication abortion have shifted during this period of increasing legal restrictions on abortion. Documenting these changes provides insight into the ways abortion care delivery systems have adapted during a period marked by both expanding telehealth opportunities and increasing legal restrictions on abortion access.

## 2. Material and Methods

### 2.1 Data Collection

We used ANSIRH’s Abortion Facility Database which provides a national census of publicly-advertising abortion facilities in the U.S., to assess trends from 2020-2025 in the number of facilities providing medication abortion including via telehealth, whether the facilities and services were brick-and-mortar or virtual, and facilities’ gestational limits for providing medication abortion. Data collection methods are described elsewhere [7,8] and include systematic web searches and mystery shopper phone calls to abortion facilities from May/June to September/October annually. Study staff conducted mystery shopper phone calls to simulate the information a potential patient would receive from calling a facility, to confirm information found on facilities’ websites, and to obtain any incomplete or unclear information. When asked about their location, callers stated they were in the local area of the clinic; if asked questions to ascertain their gestation, callers gave last menstrual period dates placing them early in the first trimester (approximately 5 weeks). Callers did not make appointments. Data on virtual clinics were collected by web searches only, since virtual clinics usually do not provide phone numbers and give comprehensive information through their websites. The University of California San Francisco Institutional Review Board approved the study.

### 2.2 Measures

To learn whether facilities offered medication abortion and until what gestation, callers asked, “What kind of abortions do you offer?” or “What kinds of abortions can I get?”, “Can I get the abortion pill?”, and “What is the limit for how far along I can be to get that type of abortion?” To assess whether the facility offered medication abortion via telehealth, callers asked “If I choose the abortion pill, would I have to come into the clinic?” and “Could I get the abortion pill mailed to me?” We counted telehealth facilities in the states they mailed medications to, and not the states where they prescribed from.

During the annual updates in 2021-2025, we coded facilities as virtual clinics if they offered abortion services via telehealth without an associated brick-and-mortar (“in-person”) facility in the same state. Virtual clinics that served patients in multiple states were counted as separate facilities, once for each state to which it mailed medications. Brick-and-mortar facilities that provided telehealth were also counted as separate facilities in each state they mailed medications to.

### 2.3 Data Analysis

In this analysis, we describe the numbers and types of medication abortion facilities and their gestational limits, comparing changes over the five-year period and documenting trends by region and state. We present frequencies on the number of facilities that offered medication abortion in general, medication abortion via telehealth, and gestational limits for those services nationally, as well as in each geographic region and state, using U.S. Census categories. We conducted these analyses using Stata 18.

## 3. Results

### 3.1 National and Regional Trends

Nearly all abortion facilities nationally provided medication abortion over the course of the study, with 99% providing in 2025 (Table 1). This trend is driven by an increase in the number and proportion of facilities that are providing only medication abortion, rising from 35% (n=264) in 2020 to 65% (n=861) in 2025 (Figure 1). These increases were seen in all regions, including those most affected by abortion bans since *Dobbs*. In the Midwest, the proportion of facilities offering medication only increased from 32% in 2020 to 67% in 2025; in the South, the increase was from 12% to 61%. In the Northeast and West, regions with greater numbers of abortion facilities, medication abortion-only service delivery increased to 62% and 69% of facilities in those regions, respectively, by 2025.

**Figure 1.**
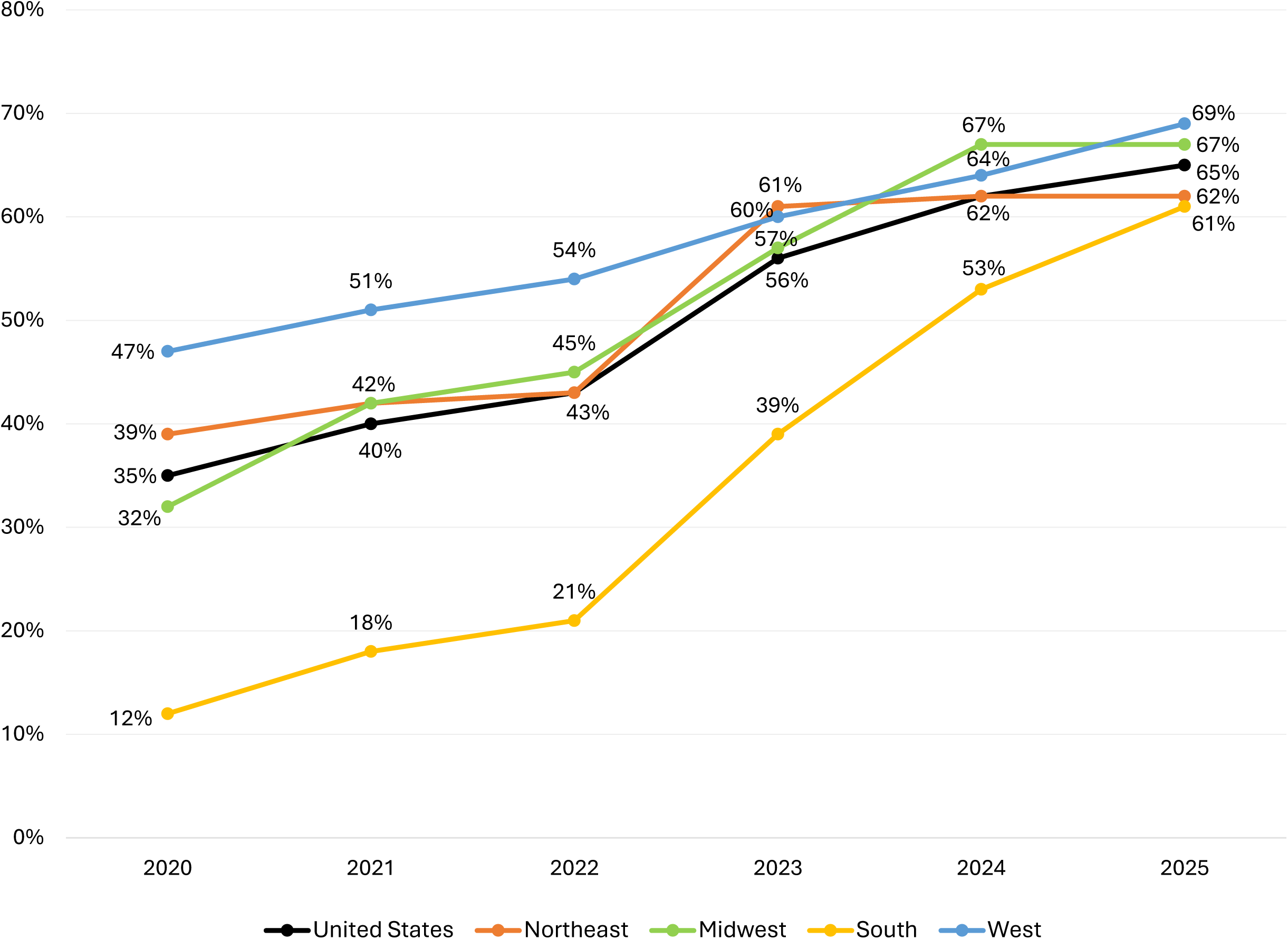
Proportion of U.S. abortion facilities providing only medication abortion, by region and year from 2020-2025.

**Table 1.** Number of U.S. abortion facilities with medication abortion services available, by geographic region and state from 2020-2025.

| Geographic Region and State | No. (%) of abortion facilities providing medication abortion |  |  |  |  |  | No. (%) of abortion facilities providing ONLY medication abortion |  |  |  |  |  |
| --- | --- | --- | --- | --- | --- | --- | --- | --- | --- | --- | --- | --- |
|  | 2020 | 2021 | 2022 | 2023 | 2024 | 2025 | 2020 | 2021 | 2022 | 2023 | 2024 | 2025 |
| **UNITED STATES (Total)** | **730 (97)** | **772 (99)** | **788 (99)** | **958 (100)** | **1,241 (100)** | **1,321 (99)** | **264 (35)** | **312 (40)** | **346 (43)** | **531 (56)** | **770 (62)** | **861 (65)** |
| **NORTHEAST** | **200 (93)** | **214 (97)** | **244 (98)** | **264 (100)** | **374 (100)** | **385 (100)** | **83 (39)** | **93 (42)** | **109 (43)** | **161 (61)** | **232 (62)** | **240 (62)** |
| **New England** | **61 (99)** | **72 (97)** | **84 (99)** | **105 (100)** | **148 (100)** | **168 (100)** | **29 (47)** | **38 (51)** | **49 (57)** | **77 (73)** | **116 (78)** | **135 (80)** |
| Connecticut | 12 (100) | 18 (100) | 22 (100) | 26 (100) | 29 (100) | 31 (100) | 6 (50) | 11 (61) | 14 (64) | 20 (77) | 22 (76) | 25 (81) |
| Maine | 20 (100) | 21 (100) | 25 (100) | 31 (100) | 35 (100) | 40 (100) | 17 (85) | 18 (86) | 21 (84) | 26 (87) | 32 (91) | 37 (92) |
| Massachusetts | 16 (94) | 16 (89) | 18 (95) | 21 (100) | 37 (100) | 37 (100) | 2 (11) | 2 (11) | 3 (15) | 11 (52) | 24 (65) | 25 (68) |
| New Hampshire | 5 (100) | 7 (100) | 7 (100) | 10 (100) | 18 (100) | 20 (100) | 1 (20) | 2 (29) | 3 (43) | 5 (50) | 12 (67) | 14 (70) |
| Rhode Island | 2 (100) | 3 (100) | 4 (100) | 8 (100) | 12 (100) | 15 (100) | 0 (0) | 1 (33) | 3 (75) | 6 (75) | 10 (83) | 1 (100) |
| Vermont | 6 (100) | 7 (100) | 8 (100) | 9 (100) | 16 (100) | 24 (100) | 3 (50) | 4 (57) | 5 (62) | 8 (89) | 15 (94) | 13 (87) |
| **Middle Atlantic** | **139 (92)** | **142 (98)** | **160 (97)** | **159 (100)** | **226 (100)** | **217 (100)** | **54 (36)** | **55 (37)** | **60 (36)** | **84 (53)** | **116 (51)** | **20 (83)** |
| New Jersey | 41 (95) | 42 (98) | 44 (98) | 47 (100) | 50 (100) | 55 (98) | 20 (47) | 20 (47) | 21 (47) | 29 (63) | 30 (60) | 32 (57) |
| New York | 83 (88) | 85 (96) | 98 (96) | 88 (98) | 149 (100) | 136 (100) | 31 (34) | 32 (36) | 33 (32) | 44 (49) | 70 (47) | 57 (42) |
| Pennsylvania | 15 (94) | 15 (100) | 18 (100) | 24 (100) | 27 (100) | 26 (100) | 3 (20) | 3 (20) | 6 (33) | 11 (48) | 16 (59) | 16 (62) |
| **MIDWEST** | **92 (99)** | **100 (98)** | **106 (100)** | **143 (100)** | **177 (100)** | **199 (99)** | **30 (32)** | **43 (42)** | **48 (45)** | **79 (57)** | **119 (67)** | **136 (67)** |
| **East North Central** | **72 (100)** | **75 (99)** | **81 (100)** | **97 (100)** | **117 (100)** | **124 (100)** | **25 (35)** | **32 (42)** | **36 (44)** | **50 (53)** | **70 (60)** | **73 (59)** |
| Illinois | 30 (100) | 29 (100) | 39 (100) | 48 (100) | 56 (100) | 55 (100) | 13 (43) | 16 (55) | 20 (53) | 26 (55) | 34 (61) | 30 (55) |
| Indiana | 7 (100) | 7 (100) | 7 (100) | 1 (100) | 4 (100) | 6 (100) | 2 (29) | 2 (29) | 3 (43) | 1 (100) | 4 (100) | 6 (100) |
| Michigan | 22 (100) | 26 (97) | 26 (100) | 33 (100) | 36 (100) | 35 (100) | 7 (32) | 11 (41) | 11 (41) | 18 (55) | 20 (56) | 19 (54) |
| Ohio | 9 (100) | 9 (100) | 9 (100) | 11 (100) | 13 (100) | 18 (100) | 2 (22) | 2 (22) | 2 (22) | 4 (36) | 7 (54) | 11 (61) |
| Wisconsin | 4 (100) | 4 (100) | -- | 4 (100) | 8 (100) | 10 (100) | 1 (25) | 1 (25) | -- | 1 (33) | 5 (62) | 7 (70) |
| **West North Central** | **20 (95)** | **25 (96)** | **25 (100)** | **46 (100)** | **60 (100)** | **75 (96)** | **5 (24)** | **11 (42)** | **12 (48)** | **29 (66)** | **49 (82)** | **63 (81)** |
| Iowa | 6 (100) | 7 (100) | 7 (100) | 9 (100) | 6 (100) | 6 (100) | 3 (50) | 5 (57) | 4 (57) | 6 (67) | 5 (83) | 6 (100) |
| Kansas | 4 (100) | 4 (100) | 4 (100) | 10 (100) | 12 (100) | 14 (100) | 1 (25) | 1 (25) | 1 (25) | 5 (50) | 8 (67) | 9 (64) |
| Minnesota | 5 (100) | 9 (100) | 10 (100) | 15 (100) | 22 (100) | 28 (100) | 1 (20) | 6 (67) | 7 (70) | 11 (73) | 18 (82) | 23 (82) |
| Missouri | x | x | -- | 2 (100) | 5 (100) | 6 (67) | x | x | -- | 2 (100) | 5 (100) | 6 (67) |
| Nebraska | 1 (100) | 1 (100) | 3 (100) | 5 (100) | 6 (100) | 8 (100) | 0 (0) | 0 (0) | 0 (0) | 3 (60) | 5 (83) | 7 (88) |
| North Dakota | 3 (100) | 3 (100) | 1 (100) | 3 (100) | 5 (100) | 7 (100) | 0 (0) | 0 (0) | 0 (0) | 1 (50) | 4 (80) | 6 (86) |
| South Dakota | 1 (100) | 1 (100) | -- | 2 (100) | 4 (100) | 6 (100) | 0 (0) | 0 (0) | -- | 1 (100) | 4 (100) | 6 (100) |
| **SOUTH** | **169 (99)** | **179 (99)** | **145 (99)** | **185 (100)** | **248 (99)** | **272 (99)** | **21 (12)** | **32 (18)** | **31 (21)** | **72 (39)** | **133 (53)** | **167 (61)** |
| **South Atlantic** | **126 (100)** | **135 (100)** | **145 (99)** | **175 (100)** | **211 (99)** | **227 (99)** | **16 (13)** | **24 (18)** | **31 (21)** | **62 (35)** | **96 (45)** | **122 (53)** |
| Delaware | 2 (100) | 3 (100) | 5 (100) | 10 (100) | 17 (100) | 18 (100) | 0 (0) | 1 (33) | 3 (60) | 7 (70) | 13 (76) | 14 (78) |
| District of Columbia | 5 (100) | 5 (100) | 8 (89) | 10 (100) | 14 (93) | 17 (94) | 0 (0) | 1 (20) | 4 (44) | 6 (60) | 11 (73) | 15 (83) |
| Florida | 52 (100) | 55 (100) | 54 (100) | 57 (100) | 54 (100) | 54 (98) | 6 (12) | 5 (9) | 4 (7) | 8 (14) | 10 (19) | 15 (27) |
| Georgia | 14 (100) | 15 (100) | 15 (100) | 17 (100) | 19 (100) | 20 (100) | 4 (29) | 6 (40) | 6 (40) | 9 (53) | 9 (47) | 11 (55) |
| Maryland | 18 (100) | 21 (100) | 24 (100) | 33 (100) | 40 (99) | 43 (100) | 4 (22) | 7 (33) | 8 (33) | 15 (47) | 23 (57) | 29 (67) |
| North Carolina | 15 (100) | 15 (94) | 16 (94) | 17 (100) | 20 (100) | 23 (100) | 1 (7) | 1 (6) | 1 (6) | 2 (12) | 5 (25) | 6 (26) |
| South Carolina | 3 (100) | 3 (100) | 3 (100) | 5 (100) | 7 (100) | 9 (100) | 0 (0) | 0 (0) | 0 (0) | 2 (40) | 4 (57) | 6 (67) |
| Virginia | 16 (100) | 17 (100) | 20 (100) | 25 (100) | 35 (100) | 37 (100) | 1 (6) | 3 (18) | 5 (25) | 11 (44) | 16 (46) | 20 (54) |
| West Virginia | 1 (100) | 1 (100) | -- | 1 (100) | 5 (100) | 6 (100) | 0 (0) | 0 (0) | -- | 1 (100) | 5 (100) | 6 (100) |
| **East South Central** | **12 (92)** | **15 (100)** | **--** | **5 (100)** | **18 (100)** | **22 (100)** | **2 (15)** | **2 (13)** | **--** | **5 (100)** | **18 (100)** | **22 (100)** |
| Alabama | 2 (67) | 5 (100) | -- | 1 (100) | 4 (100) | 5 (100) | 0 (0) | 1 (20) | -- | 1 (100) | 4 (100) | 5 (100) |
| Kentucky | 1 (100) | 2 (100) | -- | 2 (100) | 5 (100) | 6 (100) | 0 (0) | 0 (0) | -- | 2 (100) | 5 (100) | 6 (100) |
| Mississippi | 1 (100) | 1 (100) | -- | 1 (100) | 4 (100) | 5 (100) | 0 (0) | 0 (0) | -- | 1 (100) | 4 (100) | 5 (100) |
| Tennessee | 8 (100) | 7 (100) | -- | 1 (100) | 5 (100) | 6 (100) | 2 (25) | 1 (14) | -- | 1 (100) | 5 (100) | 6 (100) |
| **West South Central** | **31 (100)** | **29 (97)** | **--** | **5 (100)** | **19 (100)** | **23 (100)** | **3 (10)** | **6 (20)** | **--** | **5 (100)** | **19 (100)** | **23 (100)** |
| Arkansas | 2 (100) | 2 (100) | -- | 2 (100) | 5 (100) | 6 (100) | 1 (50) | 1 (50) | -- | 2 (100) | 5 (100) | 6 (100) |
| Louisiana | 3 (100) | 3 (100) | -- | 1 (100) | 5 (100) | 6 (100) | 0 (0) | 0 (0) | -- | 1 (100) | 5 (100) | 6 (100) |
| Oklahoma | 4 (100) | 3 (100) | -- | 1 (100) | 5 (100) | 6 (100) | 0 (0) | 0 (0) | -- | 1 (100) | 5 (100) | 5 (100) |
| Texas | 22 (100) | 21 (96) | -- | 1 (100) | 4 (100) | 5 (100) | 2 (9) | 5 (23) | -- | 1 (100) | 4 (100) | 5 (100) |
| **WEST** | **269 (98)** | **279 (99)** | **293 (99)** | **366 (100)** | **442 (99)** | **465 (100)** | **130 (47)** | **144 (51)** | **158 (54)** | **219 (60)** | **286 (64)** | **318 (69)** |
| **Mountain** | **57 (97)** | **61 (96)** | **64 (97)** | **102 (99)** | **130 (100)** | **142 (100)** | **20 (34)** | **28 (44)** | **33 (50)** | **62 (61)** | **95 (73)** | **105 (74)** |
| Arizona | 8 (100) | 8 (100) | 8 (100) | 9 (100) | 13 (100) | 15 (100) | 1 (13) | 1 (13) | 3 (38) | 3 (33) | 6 (46) | 7 (47) |
| Colorado | 22 (96) | 24 (96) | 23 (96) | 32 (97) | 40 (98) | 40 (100) | 10 (44) | 13 (52) | 11 (46) | 17 (52) | 29 (71) | 29 (72) |
| Idaho | 4 (100) | 4 (100) | -- | 2 (100) | 4 (100) | 6 (100) | 2 (50) | 2 (50) | -- | 2 (100) | 4 (100) | 6 (100) |
| Montana | 6 (100) | 7 (100) | 9 (100) | 11 (100) | 14 (100) | 16 (100) | 2 (33) | 3 (43) | 5 (56) | 8 (73) | 11 (79) | 13 (81) |
| Nevada | 8 (89) | 9 (90) | 9 (90) | 14 (100) | 18 (100) | 19 (100) | 3 (33) | 4 (44) | 5 (50) | 9 (64) | 13 (72) | 14 (74) |
| New Mexico | 5 (100) | 6 (100) | 9 (100) | 23 (100) | 28 (100) | 29 (100) | 1 (20) | 3 (50) | 5 (56) | 16 (70) | 22 (79) | 22 (76) |
| Utah | 2 (100) | 2 (100) | 4 (100) | 6 (100) | 7 (100) | 9 (100) | 0 (0) | 0 (0) | 2 (50) | 3 (60) | 5 (71) | 7 (78) |
| Wyoming | 2 (100) | 1 (100) | 2 (100) | 5 (100) | 6 (100) | 8 (100) | 1 (50) | 1 (100) | 2 (100) | 4 (80) | 5 (83) | 7 (88) |
| **Pacific** | **212 (98)** | **218 (100)** | **229 (100)** | **264 (100)** | **312 (98)** | **323 (100)** | **110 (51)** | **116 (53)** | **125 (55)** | **157 (60)** | **191 (60)** | **213 (67)** |
| Alaska | 5 (100) | 5 (100) | 4 (100) | 5 (100) | 7 (100) | 8 (100) | 1 (20) | 1 (20) | 0 (0) | 2 (40) | 4 (57) | 6 (75) |
| California | 160 (98) | 166 (100) | 174 (100) | 190 (100) | 219 (98) | 212 (100) | 88 (54) | 93 (56) | 100 (57) | 115 (61) | 132 (59) | 133 (64) |
| Hawaii | 3 (100) | 3 (100) | 3 (100) | 8 (100) | 13 (100) | 20 (100) | 0 (0) | 0 (0) | 0 (0) | 5 (63) | 10 (77) | 17 (85) |
| Oregon | 13 (100) | 14 (100) | 15 (100) | 21 (100) | 25 (100) | 29 (100) | 7 (54) | 8 (57) | 9 (60) | 13 (62) | 16 (64) | 21 (72) |
| Washington | 31 (97) | 30 (100) | 33 (100) | 40 (100) | 48 (100) | 54 (100) | 14 (44) | 14 (47) | 16 (48) | 22 (55) | 29 (60) | 36 (67) |

x Missouri did not provide medication abortion services in 2020 & 2021
-- indicates there were no documented facilities in the state/region providing abortion that year

### 3.2 Telehealth Provision

The overall increase in facilities providing medication abortion is spurred on by the growth of new facilities offering medication abortion only by telehealth, rather than a reduction of facilities offering both procedural and medication abortion. The growth occurred at both brick-and-mortar abortion facilities offering telehealth care and virtual clinics. In 2020, a small number (n=7) of brick-and-mortar facilities began to offer telehealth abortion services, which has grown steadily over the study period to 68 brick-and-mortar facilities in 2025 (Table 2, Figure 2).

**Figure 2.**
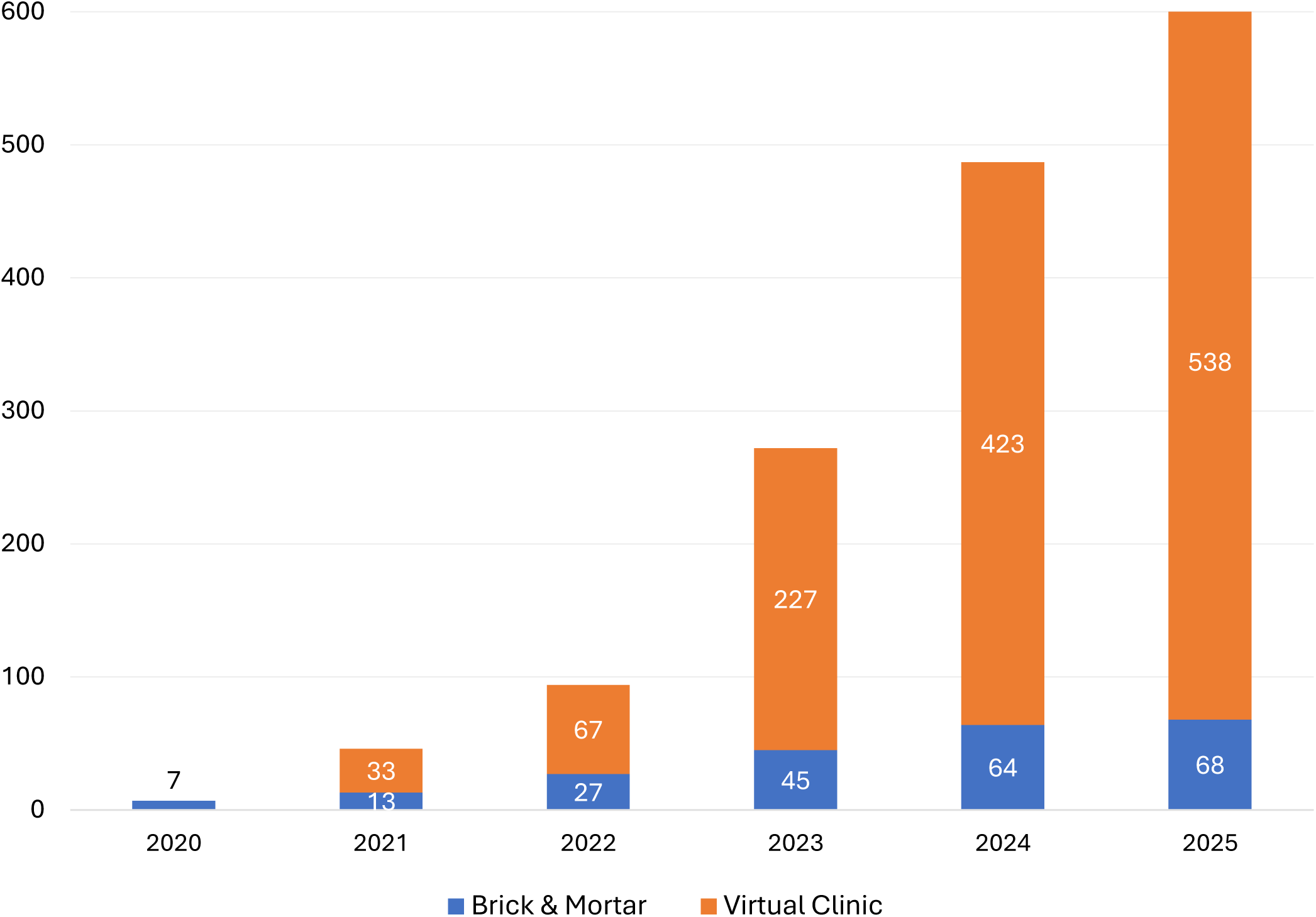
Number of U.S. abortion facilities that offer telehealth for medication abortion, by facility type and year from 2020-2025.

**Table 2.**
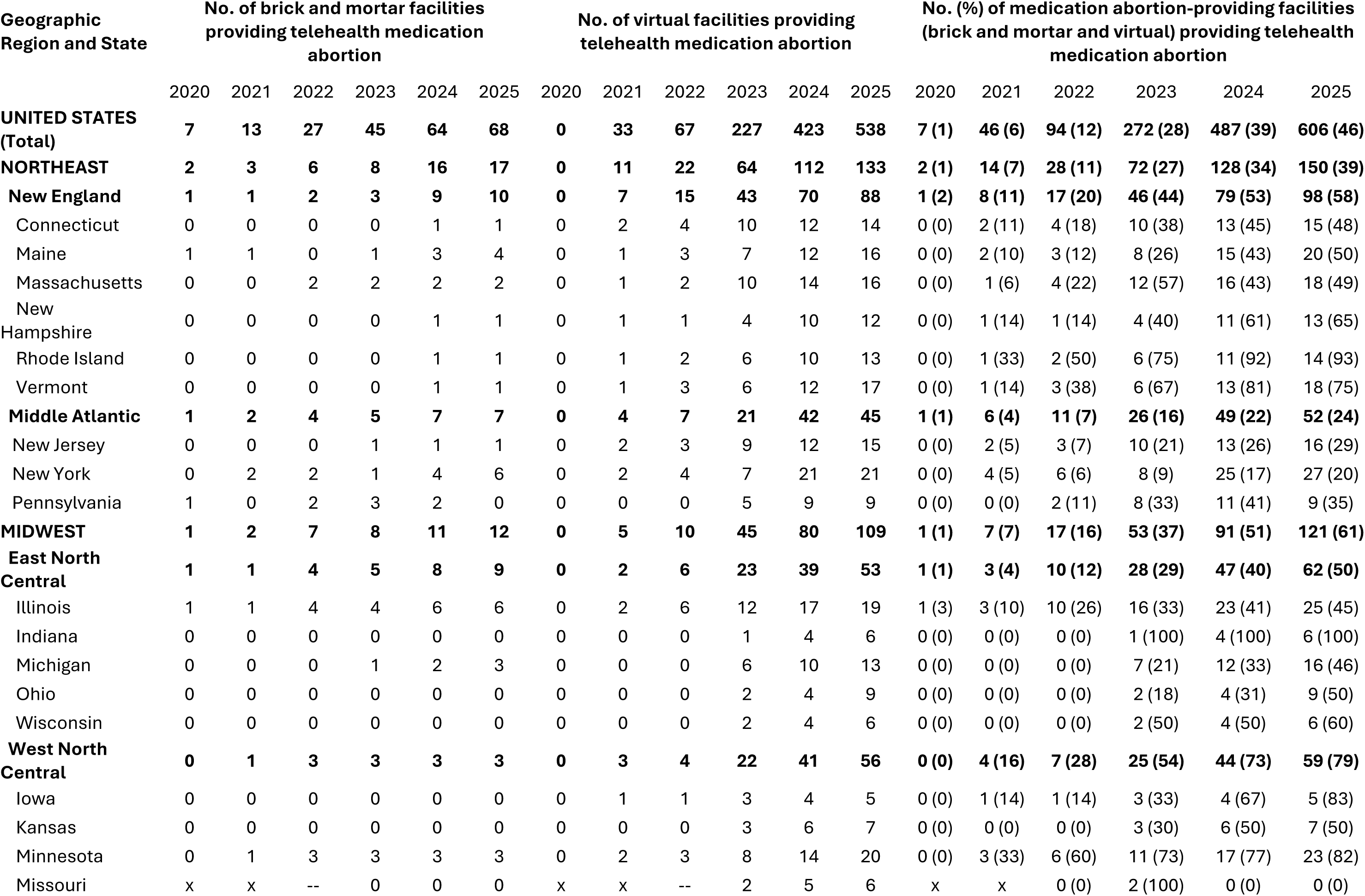
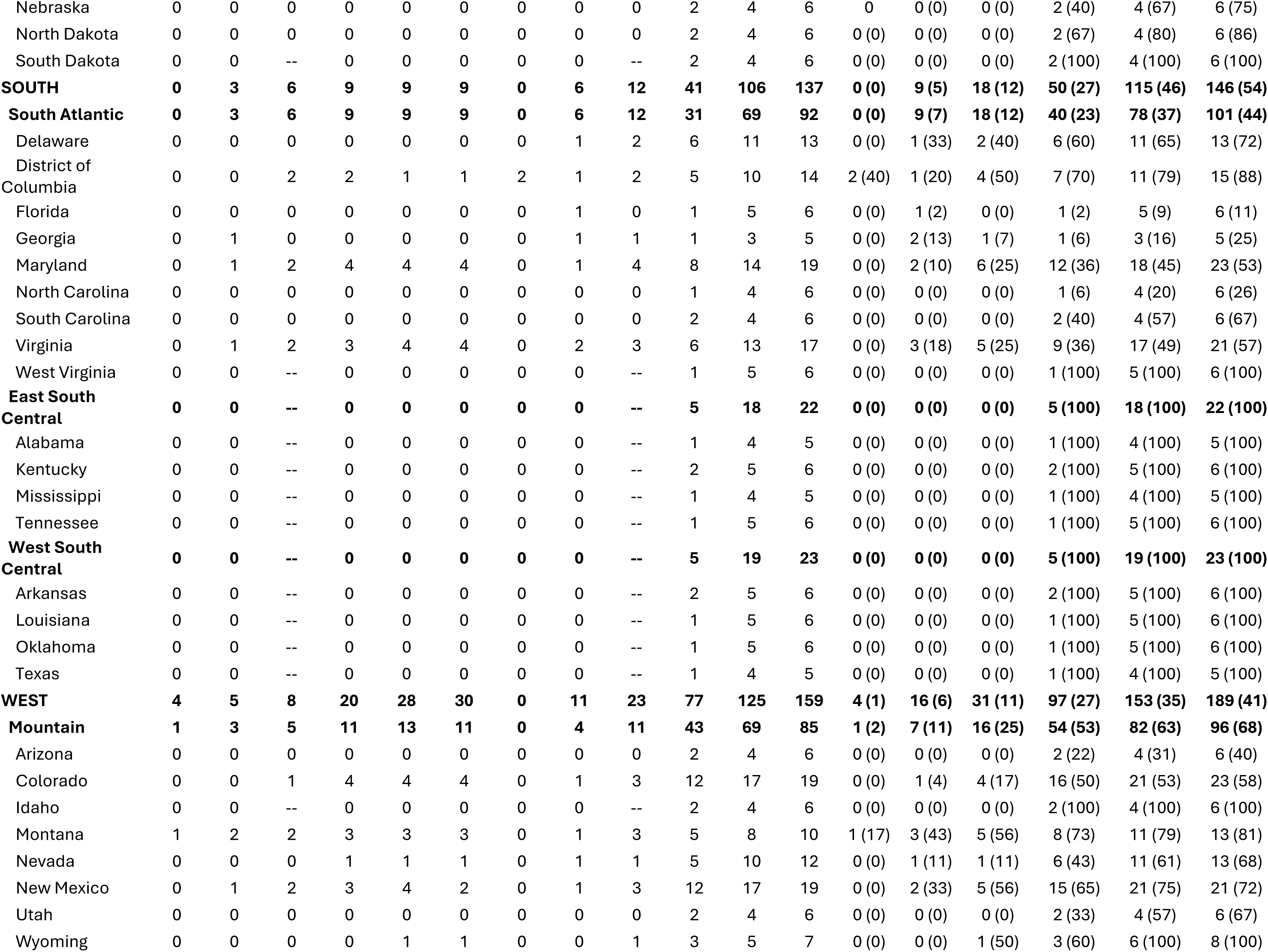
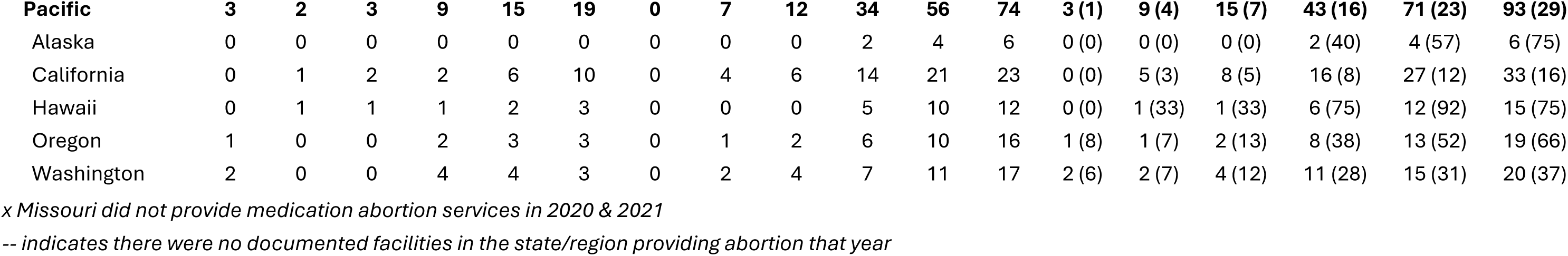
Number of U.S. abortion facilities that offer telehealth for medication abortion and facility type, by geographic region and state from 2020-2025.

| Geographic<br>Region and State | No. of brick and mortar facilities<br>providing telehealth medication<br>abortion |  |  |  |  |  | No. of virtual facilities providing<br>telehealth medication abortion |  |  |  |  |  | No. (%) of medication abortion-providing facilities<br>(brick and mortar and virtual) providing telehealth<br>medication abortion |  |  |  |  |  |
| --- | --- | --- | --- | --- | --- | --- | --- | --- | --- | --- | --- | --- | --- | --- | --- | --- | --- | --- |
|  | 2020 | 2021 | 2022 | 2023 | 2024 | 2025 | 2020 | 2021 | 2022 | 2023 | 2024 | 2025 | 2020 | 2021 | 2022 | 2023 | 2024 | 2025 |
| **UNITED STATES**<br>**(Total)** | **7** | **13** | **27** | **45** | **64** | **68** | **0** | **33** | **67** | **227** | **423** | **538** | **7 (1)** | **46 (6)** | **94 (12)** | **272 (28)** | **487 (39)** | **606 (46)** |
| **NORTHEAST** | **2** | **3** | **6** | **8** | **16** | **17** | **0** | **11** | **22** | **64** | **112** | **133** | **2 (1)** | **14 (7)** | **28 (11)** | **72 (27)** | **128 (34)** | **150 (39)** |
| **New England** | **1** | **1** | **2** | **3** | **9** | **10** | **0** | **7** | **15** | **43** | **70** | **88** | **1 (2)** | **8 (11)** | **17 (20)** | **46 (44)** | **79 (53)** | **98 (58)** |
| Connecticut | 0 | 0 | 0 | 0 | 1 | 1 | 0 | 2 | 4 | 10 | 12 | 14 | 0 (0) | 2 (11) | 4 (18) | 10 (38) | 13 (45) | 15 (48) |
| Maine | 1 | 1 | 0 | 1 | 3 | 4 | 0 | 1 | 3 | 7 | 12 | 16 | 0 (0) | 2 (10) | 3 (12) | 8 (26) | 15 (43) | 20 (50) |
| Massachusetts | 0 | 0 | 2 | 2 | 2 | 2 | 0 | 1 | 2 | 10 | 14 | 16 | 0 (0) | 1 (6) | 4 (22) | 12 (57) | 16 (43) | 18 (49) |
| New Hampshire | 0 | 0 | 0 | 0 | 1 | 1 | 0 | 1 | 1 | 4 | 10 | 12 | 0 (0) | 1 (14) | 1 (14) | 4 (40) | 11 (61) | 13 (65) |
| Rhode Island | 0 | 0 | 0 | 0 | 1 | 1 | 0 | 1 | 2 | 6 | 10 | 13 | 0 (0) | 1 (33) | 2 (50) | 6 (75) | 11 (92) | 14 (93) |
| Vermont | 0 | 0 | 0 | 0 | 1 | 1 | 0 | 1 | 3 | 6 | 12 | 17 | 0 (0) | 1 (14) | 3 (38) | 6 (67) | 13 (81) | 18 (75) |
| **Middle Atlantic** | **1** | **2** | **4** | **5** | **7** | **7** | **0** | **4** | **7** | **21** | **42** | **45** | **1 (1)** | **6 (4)** | **11 (7)** | **26 (16)** | **49 (22)** | **52 (24)** |
| New Jersey | 0 | 0 | 0 | 1 | 1 | 1 | 0 | 2 | 3 | 9 | 12 | 15 | 0 (0) | 2 (5) | 3 (7) | 10 (21) | 13 (26) | 16 (29) |
| New York | 0 | 2 | 2 | 1 | 4 | 6 | 0 | 2 | 4 | 7 | 21 | 21 | 0 (0) | 4 (5) | 6 (6) | 8 (9) | 25 (17) | 27 (20) |
| Pennsylvania | 1 | 0 | 2 | 3 | 2 | 0 | 0 | 0 | 0 | 5 | 9 | 9 | 0 (0) | 0 (0) | 2 (11) | 8 (33) | 11 (41) | 9 (35) |
| **MIDWEST** | **1** | **2** | **7** | **8** | **11** | **12** | **0** | **5** | **10** | **45** | **80** | **109** | **1 (1)** | **7 (7)** | **17 (16)** | **53 (37)** | **91 (51)** | **121 (61)** |
| **East North Central** | **1** | **1** | **4** | **5** | **8** | **9** | **0** | **2** | **6** | **23** | **39** | **53** | **1 (1)** | **3 (4)** | **10 (12)** | **28 (29)** | **47 (40)** | **62 (50)** |
| Illinois | 1 | 1 | 4 | 4 | 6 | 6 | 0 | 2 | 6 | 12 | 17 | 19 | 1 (3) | 3 (10) | 10 (26) | 16 (33) | 23 (41) | 25 (45) |
| Indiana | 0 | 0 | 0 | 0 | 0 | 0 | 0 | 0 | 0 | 1 | 4 | 6 | 0 (0) | 0 (0) | 0 (0) | 1 (100) | 4 (100) | 6 (100) |
| Michigan | 0 | 0 | 0 | 1 | 2 | 3 | 0 | 0 | 0 | 6 | 10 | 13 | 0 (0) | 0 (0) | 0 (0) | 7 (21) | 12 (33) | 16 (46) |
| Ohio | 0 | 0 | 0 | 0 | 0 | 0 | 0 | 0 | 0 | 2 | 4 | 9 | 0 (0) | 0 (0) | 0 (0) | 2 (18) | 4 (31) | 9 (50) |
| Wisconsin | 0 | 0 | 0 | 0 | 0 | 0 | 0 | 0 | 0 | 2 | 4 | 6 | 0 (0) | 0 (0) | 0 (0) | 2 (50) | 4 (50) | 6 (60) |
| **West North Central** | **0** | **1** | **3** | **3** | **3** | **3** | **0** | **3** | **4** | **22** | **41** | **56** | **0 (0)** | **4 (16)** | **7 (28)** | **25 (54)** | **44 (73)** | **59 (79)** |
| Iowa | 0 | 0 | 0 | 0 | 0 | 0 | 0 | 1 | 1 | 3 | 4 | 5 | 0 (0) | 1 (14) | 1 (14) | 3 (33) | 4 (67) | 5 (83) |
| Kansas | 0 | 0 | 0 | 0 | 0 | 0 | 0 | 0 | 0 | 3 | 6 | 7 | 0 (0) | 0 (0) | 0 (0) | 3 (30) | 6 (50) | 7 (50) |
| Minnesota | 0 | 1 | 3 | 3 | 3 | 3 | 0 | 2 | 3 | 8 | 14 | 20 | 0 (0) | 3 (33) | 6 (60) | 11 (73) | 17 (77) | 23 (82) |
| Missouri | x | x | -- | 0 | 0 | 0 | x | x | -- | 2 | 5 | 6 | x | x | 0 (0) | 2 (100) | 0 (0) | 0 (0) |
| Nebraska | 0 | 0 | 0 | 0 | 0 | 0 | 0 | 0 | 0 | 2 | 4 | 6 | 0 | 0 (0) | 0 (0) | 2 (40) | 4 (67) | 6 (75) |
| North Dakota | 0 | 0 | 0 | 0 | 0 | 0 | 0 | 0 | 0 | 2 | 4 | 6 | 0 (0) | 0 (0) | 0 (0) | 2 (67) | 4 (80) | 6 (86) |
| South Dakota | 0 | 0 | -- | 0 | 0 | 0 | 0 | 0 | -- | 2 | 4 | 6 | 0 (0) | 0 (0) | 0 (0) | 2 (100) | 4 (100) | 6 (100) |
| **SOUTH** | **0** | **3** | **6** | **9** | **9** | **9** | **0** | **6** | **12** | **41** | **106** | **137** | **0 (0)** | **9 (5)** | **18 (12)** | **50 (27)** | **115 (46)** | **146 (54)** |
| **South Atlantic** | **0** | **3** | **6** | **9** | **9** | **9** | **0** | **6** | **12** | **31** | **69** | **92** | **0 (0)** | **9 (7)** | **18 (12)** | **40 (23)** | **78 (37)** | **101 (44)** |
| Delaware | 0 | 0 | 0 | 0 | 0 | 0 | 0 | 1 | 2 | 6 | 11 | 13 | 0 (0) | 1 (33) | 2 (40) | 6 (60) | 11 (65) | 13 (72) |
| District of Columbia | 0 | 0 | 2 | 2 | 1 | 1 | 2 | 1 | 2 | 5 | 10 | 14 | 2 (40) | 1 (20) | 4 (50) | 7 (70) | 11 (79) | 15 (88) |
| Florida | 0 | 0 | 0 | 0 | 0 | 0 | 0 | 1 | 0 | 1 | 5 | 6 | 0 (0) | 1 (2) | 0 (0) | 1 (2) | 5 (9) | 6 (11) |
| Georgia | 0 | 1 | 0 | 0 | 0 | 0 | 0 | 1 | 1 | 1 | 3 | 5 | 0 (0) | 2 (13) | 1 (7) | 1 (6) | 3 (16) | 5 (25) |
| Maryland | 0 | 1 | 2 | 4 | 4 | 4 | 0 | 1 | 4 | 8 | 14 | 19 | 0 (0) | 2 (10) | 6 (25) | 12 (36) | 18 (45) | 23 (53) |
| North Carolina | 0 | 0 | 0 | 0 | 0 | 0 | 0 | 0 | 0 | 1 | 4 | 6 | 0 (0) | 0 (0) | 0 (0) | 1 (6) | 4 (20) | 6 (26) |
| South Carolina | 0 | 0 | 0 | 0 | 0 | 0 | 0 | 0 | 0 | 2 | 4 | 6 | 0 (0) | 0 (0) | 0 (0) | 2 (40) | 4 (57) | 6 (67) |
| Virginia | 0 | 1 | 2 | 3 | 4 | 4 | 0 | 2 | 3 | 6 | 13 | 17 | 0 (0) | 3 (18) | 5 (25) | 9 (36) | 17 (49) | 21 (57) |
| West Virginia | 0 | 0 | -- | 0 | 0 | 0 | 0 | 0 | -- | 1 | 5 | 6 | 0 (0) | 0 (0) | 0 (0) | 1 (100) | 5 (100) | 6 (100) |
| **East South Central** | **0** | **0** | **--** | **0** | **0** | **0** | **0** | **0** | **--** | **5** | **18** | **22** | **0 (0)** | **0 (0)** | **0 (0)** | **5 (100)** | **18 (100)** | **22 (100)** |
| Alabama | 0 | 0 | -- | 0 | 0 | 0 | 0 | 0 | -- | 1 | 4 | 5 | 0 (0) | 0 (0) | 0 (0) | 1 (100) | 4 (100) | 5 (100) |
| Kentucky | 0 | 0 | -- | 0 | 0 | 0 | 0 | 0 | -- | 2 | 5 | 6 | 0 (0) | 0 (0) | 0 (0) | 2 (100) | 5 (100) | 6 (100) |
| Mississippi | 0 | 0 | -- | 0 | 0 | 0 | 0 | 0 | -- | 1 | 4 | 5 | 0 (0) | 0 (0) | 0 (0) | 1 (100) | 4 (100) | 5 (100) |
| Tennessee | 0 | 0 | -- | 0 | 0 | 0 | 0 | 0 | -- | 1 | 5 | 6 | 0 (0) | 0 (0) | 0 (0) | 1 (100) | 5 (100) | 6 (100) |
| **West South Central** | **0** | **0** | **--** | **0** | **0** | **0** | **0** | **0** | **--** | **5** | **19** | **23** | **0 (0)** | **0 (0)** | **0 (0)** | **5 (100)** | **19 (100)** | **23 (100)** |
| Arkansas | 0 | 0 | -- | 0 | 0 | 0 | 0 | 0 | -- | 2 | 5 | 6 | 0 (0) | 0 (0) | 0 (0) | 2 (100) | 5 (100) | 6 (100) |
| Louisiana | 0 | 0 | -- | 0 | 0 | 0 | 0 | 0 | -- | 1 | 5 | 6 | 0 (0) | 0 (0) | 0 (0) | 1 (100) | 5 (100) | 6 (100) |
| Oklahoma | 0 | 0 | -- | 0 | 0 | 0 | 0 | 0 | -- | 1 | 5 | 6 | 0 (0) | 0 (0) | 0 (0) | 1 (100) | 5 (100) | 6 (100) |
| Texas | 0 | 0 | -- | 0 | 0 | 0 | 0 | 0 | -- | 1 | 4 | 5 | 0 (0) | 0 (0) | 0 (0) | 1 (100) | 4 (100) | 5 (100) |
| **WEST** | **4** | **5** | **8** | **20** | **28** | **30** | **0** | **11** | **23** | **77** | **125** | **159** | **4 (1)** | **16 (6)** | **31 (11)** | **97 (27)** | **153 (35)** | **189 (41)** |
| **Mountain** | **1** | **3** | **5** | **11** | **13** | **11** | **0** | **4** | **11** | **43** | **69** | **85** | **1 (2)** | **7 (11)** | **16 (25)** | **54 (53)** | **82 (63)** | **96 (68)** |
| Arizona | 0 | 0 | 0 | 0 | 0 | 0 | 0 | 0 | 0 | 2 | 4 | 6 | 0 (0) | 0 (0) | 0 (0) | 2 (22) | 4 (31) | 6 (40) |
| Colorado | 0 | 0 | 1 | 4 | 4 | 4 | 0 | 1 | 3 | 12 | 17 | 19 | 0 (0) | 1 (4) | 4 (17) | 16 (50) | 21 (53) | 23 (58) |
| Idaho | 0 | 0 | -- | 0 | 0 | 0 | 0 | 0 | -- | 2 | 4 | 6 | 0 (0) | 0 (0) | 0 (0) | 2 (100) | 4 (100) | 6 (100) |
| Montana | 1 | 2 | 2 | 3 | 3 | 3 | 0 | 1 | 3 | 5 | 8 | 10 | 1 (17) | 3 (43) | 5 (56) | 8 (73) | 11 (79) | 13 (81) |
| Nevada | 0 | 0 | 0 | 1 | 1 | 1 | 0 | 1 | 1 | 5 | 10 | 12 | 0 (0) | 1 (11) | 1 (11) | 6 (43) | 11 (61) | 13 (68) |
| New Mexico | 0 | 1 | 2 | 3 | 4 | 2 | 0 | 1 | 3 | 12 | 17 | 19 | 0 (0) | 2 (33) | 5 (56) | 15 (65) | 21 (75) | 21 (72) |
| Utah | 0 | 0 | 0 | 0 | 0 | 0 | 0 | 0 | 0 | 2 | 4 | 6 | 0 (0) | 0 (0) | 0 (0) | 2 (33) | 4 (57) | 6 (67) |
| Wyoming | 0 | 0 | 0 | 0 | 1 | 1 | 0 | 0 | 1 | 3 | 5 | 7 | 0 (0) | 0 (0) | 1 (50) | 3 (60) | 6 (100) | 8 (100) |
| **Pacific** | **3** | **2** | **3** | **9** | **15** | **19** | **0** | **7** | **12** | **34** | **56** | **74** | **3 (1)** | **9 (4)** | **15 (7)** | **43 (16)** | **71 (23)** | **93 (29)** |
| Alaska | 0 | 0 | 0 | 0 | 0 | 0 | 0 | 0 | 0 | 2 | 4 | 6 | 0 (0) | 0 (0) | 0 (0) | 2 (40) | 4 (57) | 6 (75) |
| California | 0 | 1 | 2 | 2 | 6 | 10 | 0 | 4 | 6 | 14 | 21 | 23 | 0 (0) | 5 (3) | 8 (5) | 16 (8) | 27 (12) | 33 (16) |
| Hawaii | 0 | 1 | 1 | 1 | 2 | 3 | 0 | 0 | 0 | 5 | 10 | 12 | 0 (0) | 1 (33) | 1 (33) | 6 (75) | 12 (92) | 15 (75) |
| Oregon | 1 | 0 | 0 | 2 | 3 | 3 | 0 | 1 | 2 | 6 | 10 | 16 | 1 (8) | 1 (7) | 2 (13) | 8 (38) | 13 (52) | 19 (66) |
| Washington | 2 | 0 | 0 | 4 | 4 | 3 | 0 | 2 | 4 | 7 | 11 | 17 | 2 (6) | 2 (7) | 4 (12) | 11 (28) | 15 (31) | 20 (37) |

*x Missouri did not provide medication abortion services in 2020 & 2021*
*-- indicates there were no documented facilities in the state/region providing abortion that year*

The study team first identified virtual clinics in web searches in 2021 after the in-person dispensing requirement for mifepristone was lifted, recording 33 virtual clinics nationally in 2021, with the number roughly doubling to 67 in 2022, then rapidly growing to 227 in 2023, 423 in 2024, and 538 in 2025. This growth was observed across regions, including states with abortion and telehealth bans, as several virtual clinics initiated services under shield law provision. In 2025, n=606 or 46% of facilities providing medication abortion (including brick-and-mortar and virtual clinics), offered telehealth medication abortion.

### 3.3 Gestational Limits

The protocols for offering medication abortion care to later pregnancy gestations expanded from 2020 to 2025. Facilities’ median gestational limits increased from 10 to 11 weeks of pregnancy nationally from 2020 to 2025, with the upper gestational limit increasing from 11 weeks to 14 weeks over that time period (Table 3). While gestational limits increased overall during the study period, some individual facilities only provided medication abortion up to 6 weeks due to 6-week abortion bans in those states (Table 3, Supplemental table). The proportion of clinics that offered medication abortion after 10 weeks increased from 32% in 2020 to 75% in 2025 and after 11 weeks increased from <1% to 38% during that time period (Figure 3). The increase in gestational limits is reflected across all geographic regions and states (Supplemental table). Those facilities offering care to later gestations tended to be virtual clinics (Figure 4), which operate under shield laws.

**Figure 3.**
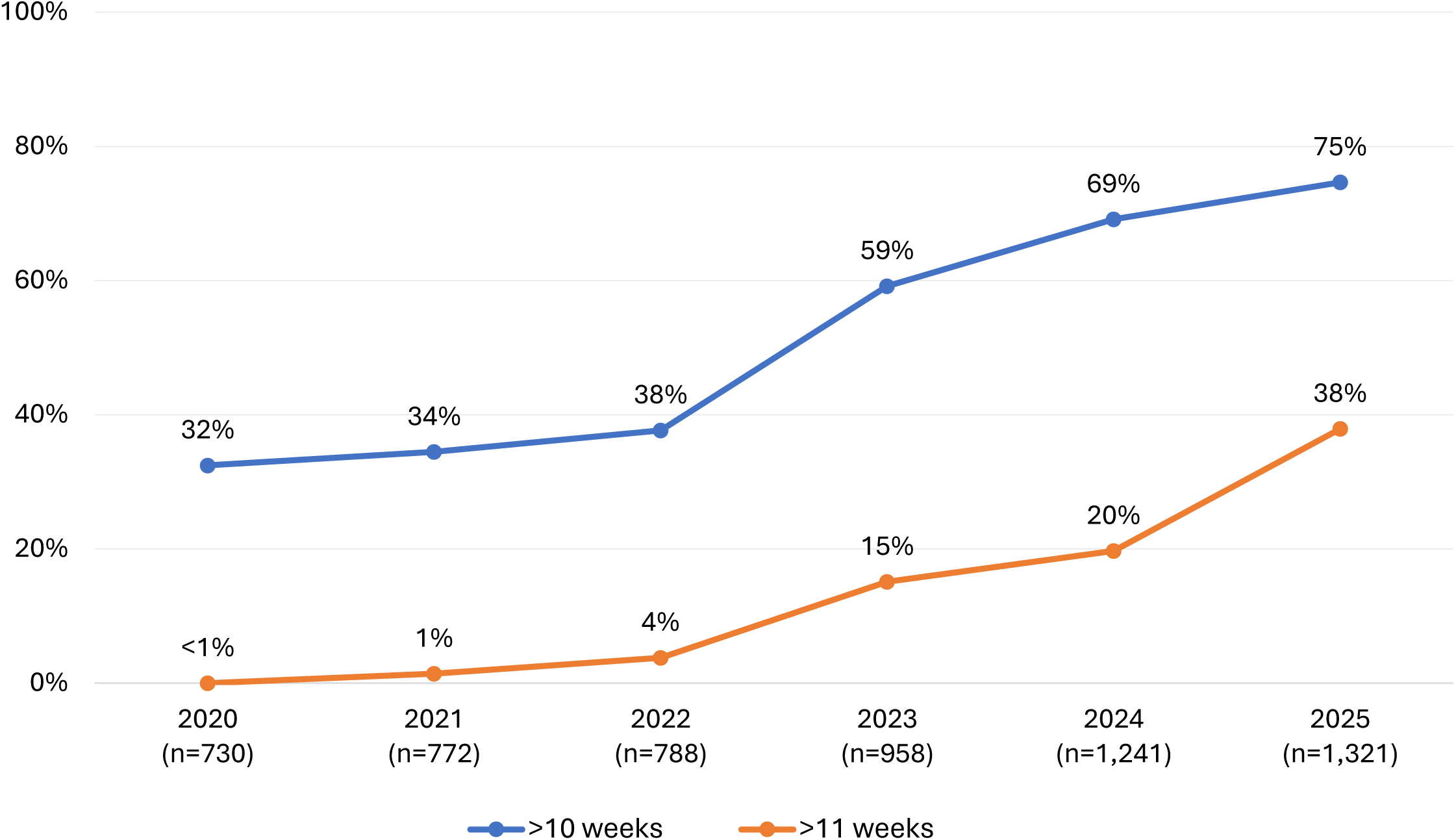
Proportion of U.S. medication abortion-providing facilities offering medication abortion after 10 and 11 weeks gestation, by year from 2020-2025.

**Figure 4.**
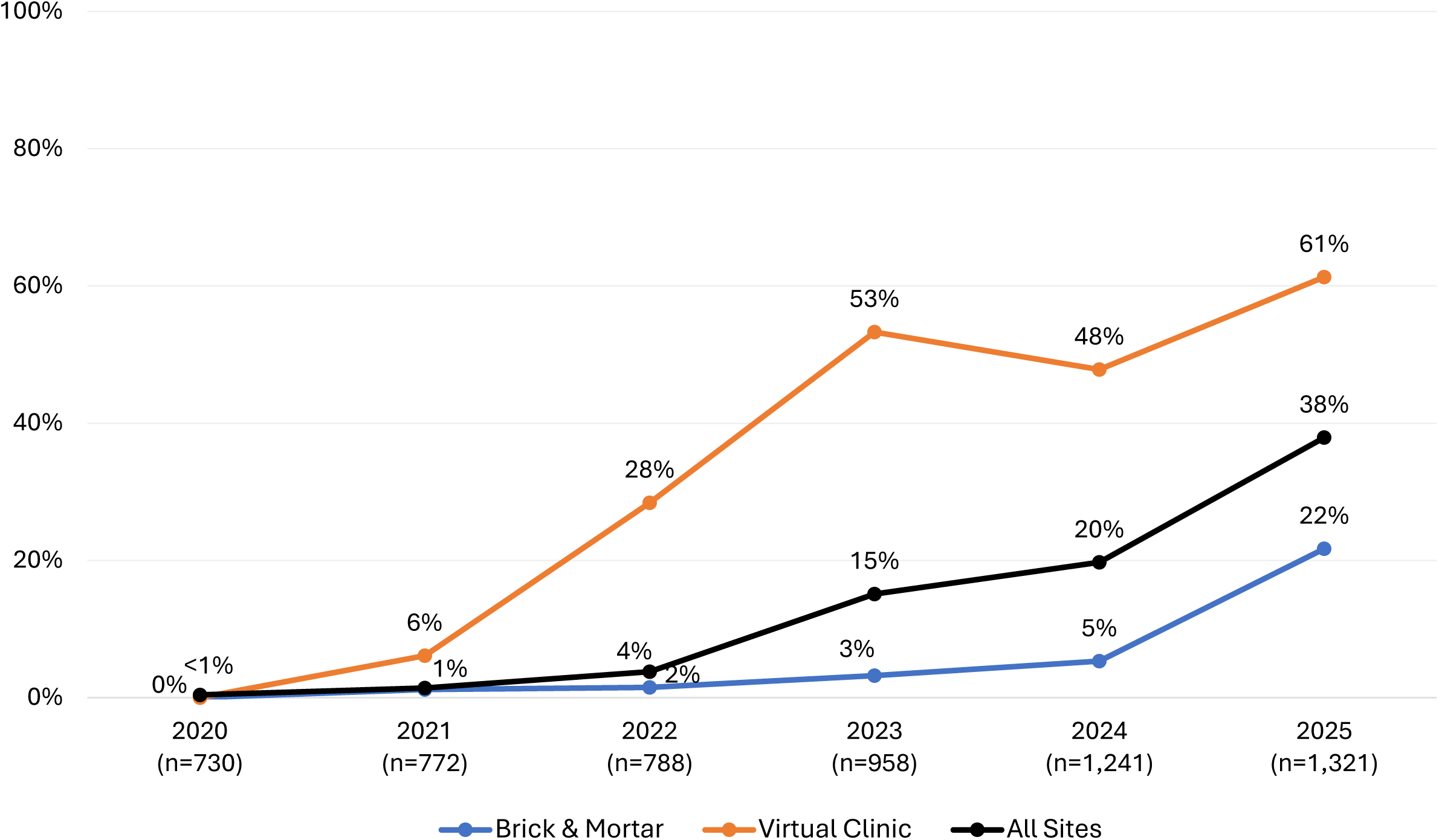
Proportion of U.S. medication abortion-providing facilities offering medication abortion after 11 weeks gestation, by facility type and year from 2020-2025. **Note**: Brick and mortar sites include all medication abortion services, which may be provided in-person or via telehealth.

**Table 3.** U.S. abortion facilities’ median gestational limits for medication abortion in weeks and facility type, by geographic region from 2020-2025.

| Geographic Region | Brick-and-mortar facilities' gestational limits for medication abortion in weeks, median (range) |  |  |  |  |  | Virtual facilities' gestational limits for medication abortion in weeks, median (range) |  |  |  |  |  | Facilities' gestational limits for medication abortion in weeks, median (range) |  |  |  |  |  |
| --- | --- | --- | --- | --- | --- | --- | --- | --- | --- | --- | --- | --- | --- | --- | --- | --- | --- | --- |
|  | 2020 | 2021 | 2022 | 2023 | 2024 | 2025 | 2020 | 2021 | 2022 | 2023 | 2024 | 2025 | 2020 | 2021 | 2022 | 2023 | 2024 | 2025 |
| **United States (Total)** | **10**<br>**(6-11)** | **10**<br>**(6-12)** | **10**<br>**(6-14)** | **11**<br>**(6-14)** | **11**<br>**(6-13)** | **11**<br>**(6-13)** | * | **8**<br>**(8-11)** | **10**<br>**(8-13)** | **12**<br>**(8-13)** | **11**<br>**(8-13)** | **12**<br>**(9-14)** | **10**<br>**(6-11)** | **10**<br>**(6-12)** | **10**<br>**(6-14)** | **11**<br>**(6-14)** | **11**<br>**(6-13)** | **11**<br>**(6-14)** |
| **NORTHEAST** | **10**<br>**(7-11)** | **10**<br>**(6-12)** | **10**<br>**(7-11)** | **11**<br>**(7-12)** | **11**<br>**(7-12)** | **11**<br>**(7-13)** | * | **8**<br>**(8-11)** | **10**<br>**(8-13)** | **11**<br>**(8-13)** | **11**<br>**(8-13)** | **12**<br>**(10-14)** | **10**<br>**(7-11)** | **10**<br>**(6-12)** | **10**<br>**(7-13)** | **11**<br>**(7-13)** | **11**<br>**(7-13)** | **11**<br>**(7-14)** |
| New England | 10<br>(8-11) | 10<br>(8-12) | 10<br>(8-11) | 11<br>(7-12) | 11<br>(9-12) | 11<br>(9-13) | * | 8<br>(8-11) | 10<br>(8-13) | 11<br>(8-13) | 11<br>(8-13) | 12<br>(10-14) | 10<br>(8-11) | 10<br>(8-12) | 10<br>(8-13) | 11<br>(7-13) | 11<br>(8-13) | 11<br>(9-14) |
| Middle Atlantic | 11<br>(7-11) | 10<br>(6-12) | 10<br>(7-11) | 11<br>(7-12) | 11<br>(7-12) | 11<br>(7-13) | * | 9<br>(8-11) | 10<br>(8-13) | 11<br>(9-13) | 11<br>(9-13) | 11<br>(10-14) | 11<br>(7-11) | 10<br>(6-12) | 10<br>(7-13) | 11<br>(7-13) | 11<br>(7-13) | 11<br>(7-14) |
| **MIDWEST** | **10**<br>**(9-11)** | **10**<br>**(6-11)** | **11**<br>**(6-11)** | **11**<br>**(9-11)** | **11**<br>**(6-12)** | **11**<br>**(6-13)** | * | **10**<br>**(8-11)** | **10**<br>**(8-13)** | **12**<br>**(10-13)** | **12**<br>**(10-13)** | **12**<br>**(10-14)** | **10**<br>**(9-11)** | **10**<br>**(6-11)** | **11**<br>**(6-13)** | **11**<br>**(9-13)** | **11**<br>**(6-13)** | **12**<br>**(6-14)** |
| East North Central | 10<br>(9-11) | 10<br>(6-11) | 10<br>(6-11) | 11<br>(9-11) | 11<br>(6-12) | 11<br>(9-13) | * | 9<br>(8-10) | 10<br>(8-13) | 12<br>(10-13) | 12<br>(10-13) | 12<br>(10-14) | 10<br>(9-11) | 10<br>(6-11) | 10<br>(6-13) | 11<br>(9-13) | 11<br>(6-13) | 12<br>(9-14) |
| West North Central | 10<br>(9-11) | 11<br>(10-11) | 11<br>(10-11) | 11<br>(9-11) | 11<br>(6-12) | 11<br>(6-12) | * | 10<br>(8-11) | 10<br>(8-11) | 12<br>(10-13) | 13<br>(10-13) | 13<br>(10-14) | 10<br>(9-11) | 11<br>(8-11) | 11<br>(8-11) | 11<br>(9-13) | 11<br>(6-13) | 12<br>(6-14) |
| **SOUTH** | **10**<br>**(6-11)** | **10**<br>**(6-12)** | **10**<br>**(6-14)** | **10**<br>**(6-14)** | **8**<br>**(6-13)** | **8**<br>**(6-13)** | * | **8**<br>**(8-11)** | **10**<br>**(8-13)** | **12**<br>**(10-13)** | **12**<br>**(10-13)** | **13**<br>**(10-14)** | **10**<br>**(6-11)** | **10**<br>**(6-12)** | **10**<br>**(6-14)** | **11**<br>**(6-14)** | **11**<br>**(6-13)** | **11**<br>**(6-14)** |
| South Atlantic | 10<br>(6-11) | 10<br>(8-12) | 10<br>(6-14) | 10<br>(6-14) | 8<br>(6-13) | 8<br>(6-13) | * | 8<br>(8-11) | 10<br>(8-13) | 12<br>(10-13) | 12<br>(10-13) | 12<br>(10-14) | 10<br>(6-11) | 10<br>(6-12) | 10<br>(6-14) | 11<br>(6-14) | 11<br>(6-13) | 11<br>(6-14) |
| East South Central | 11<br>(8-11) | 11<br>(8-12) | -- | -- | -- | -- | * | * | -- | 12<br>(12-13) | 12<br>(11-13) | 13<br>(11-14) | 11<br>(8-11) | 11<br>(8-12) | -- | 12<br>(12-13) | 12<br>(11-13) | 13<br>(11-14) |
| West South Central | 10<br>(6-11) | 6<br>(6-11) | -- | -- | -- | -- | * | * | -- | 12<br>(12-13) | 13<br>(11-13) | 13<br>(11-14) | 10<br>(6-11) | 6<br>(6-11) | -- | 12<br>(12-13) | 13<br>(11-13) | 13<br>(11-14) |
| **WEST** | **10**<br>**(7-11)** | **10**<br>**(7-12)** | **10**<br>**(7-12)** | **10**<br>**(7-13)** | **11**<br>**(6-12)** | **11**<br>**(6-12)** | * | **8**<br>**(8-10)** | **10**<br>**(8-13)** | **11**<br>**(10-13)** | **11**<br>**(9-13)** | **12**<br>**(9-14)** | **10**<br>**(7-11)** | **10**<br>**(7-12)** | **10**<br>**(7-13)** | **11**<br>**(7-13)** | **11**<br>**(6-13)** | **11**<br>**(6-14)** |
| Mountain | 10<br>(7-11) | 11<br>(7-12) | 11<br>(7-12) | 11<br>(8-13) | 11<br>(8-12) | 11<br>(8-12) | * | 8<br>(8-8) | 10<br>(8-13) | 11<br>(10-13) | 11<br>(10-13) | 12<br>(10-14) | 10<br>(7-11) | 10<br>(7-12) | 11<br>(7-13) | 11<br>(8-13) | 11<br>(8-13) | 11<br>(8-14) |
| Pacific | 10<br>(7-11) | 10<br>(7-12) | 10<br>(7-12) | 10<br>(7-11) | 11<br>(6-12) | 11<br>(6-12) | * | 10<br>(8-10) | 10<br>(8-13) | 11<br>(10-13) | 11<br>(9-13) | 11<br>(9-14) | 10<br>(7-11) | 10<br>(7-12) | 10<br>(7-13) | 10<br>(7-13) | 11<br>(6-13) | 11<br>(6-14) |

-- indicates there were no documented facilities in the region providing abortion that year
\* indicates there were no documented virtual/telehealth facilities providing medication abortion in the region that year

## 4. Discussion

Starting with the COVID-19 pandemic and continuing after the *Dobbs* decision, we found increasing numbers of medication abortion-only facilities, including many new virtual clinics. Abortion facilities also adapted their services to include telehealth options and extend gestational limits for medication abortion, expanding the eligibility window for people facing increased barriers to care. More facilities offering medication abortion expands outlets for abortion access, particularly with telehealth options that reduce the need to travel for care which can be logistically and financially burdensome [9–11]. These changes in service provision may help explain why the majority of all abortions are now medication abortions [1]. Taken together, these findings demonstrate substantial changes in medication abortion service delivery during a period of major policy and regulatory change.

From 2020 to 2025, we also found an increase in facilities offering medication abortion after 10 and 11 weeks. As more states enacted abortion bans following *Dobbs*, some facilities may have increased their gestational limits for medication abortion to accommodate patients experiencing delays in care because of increased travel burdens and other barriers [12]. State Medicaid policies on gestational limits for medication abortion affect reimbursements, however, and may prevent or aid facilities’ ability to provide the service at later gestations. The state Medicaid program in California, for example, recently expanded access with a bill to reimburse providers for medication abortions up until 77 days (11 weeks) gestation [13]. Other factors like provider training, provider comfort, and facility practices and costs may also affect a facility’s gestational limits for medication abortion [14]. Such increases in access to medication abortion, however, do little for patients who are contraindicated for medication abortion, including those later in pregnancy, and cannot fully substitute for access to procedural abortion care.

A key strength of this study is its use of a longitudinal census of publicly advertising abortion facilities that provides national data, detailed by region and state, to observe trends over time in medication abortion service delivery, including virtual care, telehealth care provided by brick-and-mortar facilities, and gestational limits. A limitation of these data is that they provide annual snapshots of abortion service delivery in constantly changing policy settings. The accuracy of the data is dependent on up-to-date websites and information given by clinic staff over the phone. Another consideration is that the database captures only abortion services that are publicly advertised on websites during the annual database update.

Taken together, the study findings reflect how abortion facilities, including longstanding brick-and-mortar facilities and new virtual clinics, are adapting to the evolving abortion care landscape to meet patients’ needs by increasing medication abortion service availability and providing medication abortion at later gestations. While studies show that telehealth abortion is safe and effective up to 11 weeks [15], and a few studies demonstrate medication abortion is safe and effective up to 16 weeks [16,17], further research on the safety and efficacy of medication abortion up to 14-16 weeks can support further expansion of this care. The repeal of unfounded telehealth bans and in-person visit requirements in nearly half of states and territories [18] could greatly improve medication abortion access in those areas. Protective legislation like shield laws can also support medication abortion care and thwart legal attacks that threaten this growing structure of care.

In conclusion, medication abortion service delivery has adapted to legal and logistical challenges by increasing outlets where medication abortion is available, adding telehealth options, including virtual care, and expanding gestational limits. As legal challenges, telehealth restrictions, and other barriers continue to threaten abortion access, sustained efforts by providers, researchers, and policymakers will be essential to protect, strengthen, and expand innovative models of medication abortion care.

## Data Availability

Data from ANISRH Abortion Facility Database are available upon request and approval.

https://abortionfacilitydatabase-ucsf.hub.arcgis.com/

## Acknowledgements

The authors thank the interns who assisted with data collection, and abortion facility staff for taking time to share information about their services.

## Funding

The study was supported by a core funding grant from the Advancing New Standards in Reproductive Health Program, grant # A146416, which is supported by an anonymous private foundation. Aside from this funding, authors and their institutions did not receive payment or services from a third party for any aspect of the submitted work.

## Notes

### Competing Interest Statement

The authors have declared no competing interest.

### Author Declarations

The Institutional Review Board of University of California, San Francisco gave ethical approval for this work.

