## Supplemental table for "Trends in medication abortion service delivery in the U.S., 2020-2025"

**Supplemental table. U.S. abortion facilities' median gestational limits for medication abortion in weeks, by geographic region and state from 2020-2025**

| **Geographic Region  and State** | **Facilities' gestational limits for medication abortion in weeks, median (range)** | | | | | |
| --- | --- | --- | --- | --- | --- | --- |
|  | 2020 | 2021 | 2022 | 2023 | 2024 | 2025 |
| **UNITED STATES (Total)** | **10 (6-11)** | **10 (6-12)** | **10 (6-14)** | **11 (6-14)** | **11 (6-13)** | **11 (6-14)** |
| **NORTHEAST** | **10 (7-11)** | **10 (6-12)** | **10 (7-13)** | **11 (7-13)** | **11 (7-13)** | **11 (7-14)** |
| **New England** | **10 (8-11)** | **10 (8-12)** | **10 (8-13)** | **11 (7-13)** | **11 (8-13)** | **11 (9-14)** |
| Connecticut | 11 (10-11) | 10 (8-11) | 10 (8-13) | 10 (10-13) | 11 (10-13) | 11 (9-14) |
| Maine | 10 (10-11) | 10 (8-11) | 10 (8-13) | 11 (10-13) | 11 (10-13) | 11 (10-14) |
| Massachusetts | 10 (8-11) | 10 (8-11) | 10 (8-13) | 11 (8-13) | 11 (8-13) | 11 (10-14) |
| New Hampshire | 10 (9-11) | 9 (8-11) | 9 (8-11) | 11 (7-13) | 11 (9-13) | 11 (9-14) |
| Rhode Island | 10 (10-10) | 10 (8-10) | 10 (8-13) | 11 (10-13) | 11 (10-13) | 12 (10-14) |
| Vermont | 11 (11-11) | 11 (8-12) | 11 (9-13) | 11 (10-13) | 11 (10-13) | 12 (10-14) |
| **Middle Atlantic** | **11 (7-11)** | **10 (6-12)** | **10 (7-13)** | **11 (7-13)** | **11 (7-13)** | **11 (7-14)** |
| New Jersey | 10 (7-11) | 10 (7-11) | 10 (7-13) | 10 (7-13) | 10 (8-13) | 11 (7-14) |
| New York | 11 (7-11) | 10 (6-12) | 10 (7-13) | 11 (7-13) | 11 (7-13) | 11 (7-14) |
| Pennsylvania | 11 (10-11) | 11 (10-11) | 11 (10-11) | 11 (9-13) | 11 (10-13) | 11 (10-14) |
| **MIDWEST** | **10 (9-11)** | **10 (6-11)** | **11 (6-13)** | **11 (9-13)** | **11 (6-13)** | **12 (6-14)** |
| **East North Central** | **10 (9-11)** | **10 (6-11)** | **10 (6-13)** | **11 (9-13)** | **11 (6-13)** | **12 (9-14)** |
| Illinois | 10 (10-11) | 10 (8-11) | 11 (8-13) | 11 (9-13) | 11 (6-13) | 11 (10-14) |
| Indiana | 10 (10-10) | 10 (10-10) | 10 (10-10) | 12 (12-12) | 13 (11-13) | 13 (11-14) |
| Michigan | 10 (9-11) | 10 (6-11) | 10 (6-11) | 11 (9-13) | 11 (10-13) | 12 (10-14) |
| Ohio | 10 (10-10) | 10 (10-10) | 10 (6-10) | 10 (10-13) | 10 (10-13) | 12 (9-14) |
| Wisconsin | 11 (11-11) | 11 (11-11) | -- | 12 (11-13) | 11 (11-13) | 12 (11-14) |
| **West North Central** | **10 (9-11)** | **11 (8-11)** | **11 (8-11)** | **11 (9-13)** | **11 (6-13)** | **12 (6-14)** |
| Iowa | 10 (10-11) | 11 (11-11) | 11 (10-11) | 11 (10-13) | 12 (6-13) | 13 (6-14) |
| Kansas | 10 (10-11) | 11 (10-11) | 11 (10-11) | 11 (10-12) | 11 (10-13) | 12 (10-14) |
| Minnesota | 10 (9-11) | 11 (8-11) | 11 (8-11) | 11 (9-13) | 11 (10-13) | 11 (10-14) |
| Missouri | x | x | -- | 12 (12-13) | 13 (11-13) | 13 (11-14) |
| Nebraska | 10 (10-11) | 11 (11-11) | 11 (11-11) | 11 (11-13) | 12 (11-13) | 12 (11-14) |
| North Dakota | 10 (10-10) | 10 (10-10) | 10 (10-10) | 12 (10-13) | 13 (11-13) | 13 (11-14) |
| South Dakota | 10 (10-10) | 10 (10-10) | -- | 12 (12-13) | 12 (11-13) | 13 (11-14) |
| **SOUTH** | **10 (6-11)** | **10 (6-12)** | **10 (6-14)** | **11 (6-14)** | **11 (6-13)** | **11 (6-14)** |
| **South Atlantic** | **10 (6-11)** | **10 (6-12)** | **10 (6-14)** | **11 (6-14)** | **11 (6-13)** | **11 (6-14)** |
| Delaware | 11 (11-11) | 10 (8-10) | 10 (8-13) | 11 (10-13) | 11 (10-13) | 11 (10-14) |
| District of Columbia | 10 (9-11) | 10 (8-10) | 10 (8-13) | 11 (10-13) | 11 (10-13) | 11 (10-14) |
| Florida | 10 (6-11) | 10 (6-12) | 10 (6-14) | 10 (7-14) | 6 (6-13) | 6 (6-14) |
| Georgia | 11 (8-11) | 11 (8-11) | 10 (6-11) | 6 (6-12) | 6 (6-13) | 6 (6-14) |
| Maryland | 10 (8-11) | 10 (8-11) | 10 (8-11) | 11 (9-13) | 11 (9-13) | 11 (9-14) |
| North Carolina | 10 (9-11) | 11 (8-11) | 11 (8-11) | 11 (9-12) | 11 (6-13) | 12 (9-14) |
| South Carolina | 11 (10-11) | 11 (10-11) | 11 (10-11) | 11 (6-13) | 11 (6-13) | 12 (6-14) |
| Virginia | 10 (8-11) | 10 (8-11) | 10 (8-13) | 11 (8-13) | 11 (8-13) | 11 (8-14) |
| West Virginia | 11 (11-11) | 11 (11-11) | -- | 12 (12-12) | 13 (11-13) | 13 (11-14) |
| **East South Central** | **11 (8-11)** | **11 (8-12)** | **--** | **12 (12-13)** | **12 (11-13)** | **13 (11-14)** |
| Alabama | 10 (9-10) | 11 (9-12) | -- | 12 (12-12) | 12 (11-13) | 13 (11-14) |
| Kentucky | 10 (10-10) | 10 (10-10) | -- | 12 (12-13) | 13 (11-13) | 13 (11-14) |
| Mississippi | 11 (11-11) | 11 (11-11) | -- | 12 (12-12) | 12 (11-13) | 13 (11-14) |
| Tennessee | 11 (8-11) | 11 (8-11) | -- | 12 (12-12) | 12 (11-13) | 13 (11-14) |
| **West South Central** | **10 (6-11)** | **6 (6-11)** | **--** | **12 (12-13)** | **13 (11-13)** | **13 (11-14)** |
| Arkansas | 10 (10-11) | 10 (10-10) | -- | 12 (12-13) | 13 (11-13) | 13 (11-14) |
| Louisiana | 9 (8-10) | 9 (8-10) | -- | 12 (12-12) | 13 (11-13) | 13 (11-14) |
| Oklahoma | 9 (6-11) | 10 (10-11) | -- | 12 (12-12) | 13 (11-13) | 13 (11-14) |
| Texas | 10 (9-11) | 6 (6-6) | -- | 12 (12-12) | 12 (11-13) | 13 (11-14) |
| **WEST** | **10 (7-11)** | **10 (7-12)** | **10 (7-13)** | **11 (7-13)** | **11 (6-13)** | **11 (6-14)** |
| **Mountain** | **10 (7-11)** | **10 (7-12)** | **11 (7-13)** | **11 (8-13)** | **11 (8-13)** | **11 (8-14)** |
| Arizona | 10 (10-11) | 10 (8-11) | 10 (10-11) | 11 (10-13) | 11 (10-13) | 11 (11-14) |
| Colorado | 11 (7-11) | 11 (7-11) | 11 (7-13) | 11 (8-13) | 11 (8-13) | 11 (8-14) |
| Idaho | 11 (8-11) | 10 (8-11) | -- | 12 (12-13) | 13 (11-13) | 13 (11-14) |
| Montana | 10 (10-10) | 11 (8-12) | 11 (8-13) | 11 (10-13) | 11 (10-13) | 11 (10-14) |
| Nevada | 10 (8-10) | 10 (8-11) | 10 (8-11) | 10 (8-13) | 11 (8-13) | 11 (8-14) |
| New Mexico | 11 (10-11) | 11 (8-11) | 11 (8-13) | 11 (10-13) | 11 (10-13) | 11 (10-14) |
| Utah | 9 (8-10) | 10 (9-10) | 11 (9-11) | 11 (9-13) | 11 (9-13) | 12 (9-14) |
| Wyoming | 10 (10-10) | 10 (10-10) | 10 (10-10) | 11 (10-13) | 12 (10-13) | 12 (10-14) |
| **Pacific** | **10 (7-11)** | **10 (7-12)** | **10 (7-13)** | **10 (7-13)** | **11 (6-13)** | **11 (6-14)** |
| Alaska | 11 (10-11) | 11 (10-11) | 11 (7-11) | 11 (11-13) | 11 (11-13) | 12 (11-14) |
| California | 10 (7-11) | 10 (7-11) | 10 (7-13) | 10 (7-13) | 11 (6-13) | 11 (6-14) |
| Hawaii | 11 (10-11) | 11 (11-11) | 11 (11-11) | 11 (10-13) | 11 (10-13) | 11 (10-14) |
| Oregon | 10 (8-11) | 11 (8-12) | 11 (8-13) | 11 (8-13) | 11 (10-13) | 11 (9-14) |
| Washington | 11 (10-11) | 11 (8-11) | 11 (8-13) | 11 (10-13) | 11 (10-13) | 11 (10-14) |

*x Missouri did not provide medication abortion services in 2020 & 2021*

*-- indicates there were no documented facilities in the state/region providing abortion that year*
